# Intermuscular Adiposity is Associated with Coronary Microvascular Dysfunction Independently of Body Mass Index and Modifies its Effect on Adverse Cardiovascular Outcomes

**DOI:** 10.1101/2023.05.03.23289408

**Authors:** Ana C. Souza, Troschel Amelie S., Marquardt Jan P., Moura Filipe A., Divakaran Sanjay, Hainer Jon, Blankstein Ron, Dorbala Sharmila, Di Carli Marcelo F., Fintelmann Florian J., Taqueti Viviany R.

## Abstract

**Background:** Skeletal muscle (SM) fat infiltration, or intermuscular adipose tissue (IMAT), reflects muscle quality and is associated with inflammation, a key determinant in cardiometabolic disease. Coronary flow reserve (CFR), a marker of coronary microvascular dysfunction (CMD), is independently associated with BMI, inflammation and risk of heart failure, myocardial infarction and death. We sought to investigate the relationship between skeletal muscle quality, CMD and cardiovascular outcomes.

**Methods:** Consecutive patients (N=669) undergoing evaluation for CAD with cardiac stress PET demonstrating normal perfusion and preserved left ventricular ejection fraction were followed over median 6 years for major adverse cardiovascular events (MACE), including death and hospitalization for myocardial infarction or heart failure. CFR was calculated as stress/rest myocardial blood flow and CMD defined as CFR<2. Subcutaneous adipose tissue (SAT), SM and IMAT areas (cm^2^) were obtained from simultaneous PET attenuation correction CTs using semi-automated segmentation at the twelfth thoracic vertebra (T12) level.

**Results:** Median age was 63 years, 70% were female and 46% nonwhite. Nearly half of patients were obese (46%, BMI 30-61) and BMI correlated highly with SAT and IMAT (r=0.84 and 0.71, respectively, p<0.001) and moderately with SM (r=0.52, p<0.001). Decreased SM and increased IMAT, but not BMI or SAT, remained independently associated with decreased CFR (adjusted p=0.03 and p=0.04, respectively). In adjusted analyses, both lower CFR and higher IMAT were associated with increased MACE [HR 1.78 (1.23-2.58) per -1U CFR and 1.53 (1.30-1.80) per +10 cm^2^ IMAT, adjusted p=0.002 and p<0.0001, respectively], while higher SM and SAT were protective [HR 0.89 (0.81-0.97) per +10 cm^2^ SM and 0.94 (0.91-0.98) per +10 cm^2^ SAT, adjusted p=0.01 and 0.003, respectively]. Every 1% increase in fatty muscle fraction [IMAT/(SM+IMAT)] conferred an independent 2% increased odds of CMD [CFR<2, OR 1.02 (1.01-1.04), adjusted p=0.04] and a 7% increased risk of MACE [HR 1.07 (1.04-1.09), adjusted p<0.001]. There was a significant interaction between CFR and IMAT, not BMI, such that patients with both CMD and fatty muscle demonstrated highest MACE risk (adjusted p=0.02).

**Conclusion:** Increased intermuscular fat is associated with CMD and adverse cardiovascular outcomes independently of BMI and conventional risk factors. The presence of CMD and skeletal muscle fat infiltration identified a novel at-risk cardiometabolic phenotype.

## Introduction

Overweight or obesity is prevalent in over 71% of US adults^1^ and has become one of the most important threats to public health worldwide. Compared to individuals with normal weight, those with obesity, defined as a body mass index (BMI) ≥30 kg/m^2^, experience cardiovascular disease (CVD) events at an earlier age and have a shorter average life span.^2, 3^ Excess adiposity accelerates atherosclerosis and promotes adverse changes in cardiac structure and function through deleterious effects on the myocardium as well as the vasculature, and through obesity-related comorbidities, including hypertension, dyslipidemia and type 2 diabetes mellitus.^4, 5^ Although increasing BMI is associated with increasing risk of morbidity and mortality across populations, CVD risk is not uniform for individuals with similar BMI and can vary substantially across sex and racial/ethnic groups. Nonetheless, BMI thresholds continue to guide current clinical diagnosis of obesity and candidacy for interventions with potential to improve CVD outcomes. Besides BMI, other discriminators of cardiovascular risk are needed in individuals at risk for cardiometabolic disease. Imaging technologies such as computed tomography (CT) can be used to assess body composition and distinguish between fat and lean mass directly in vivo via their respective radiodensities, or attenuation, within anatomical compartments.^6^

Recently, intermuscular adipose tissue (IMAT) has emerged as a distinct adipose depot reflecting SM fat infiltration with unique and evolving biological properties. Whereas fatty ‘marbling’ of meat is commercially valued in livestock, IMAT in humans has been associated with insulin resistance and type 2 diabetes.^7, 8^ IMAT can be found in most skeletal muscle groups, and while IMAT increases with BMI, it can vary considerably between individuals. Early reports suggest that IMAT has a proinflammatory secretome with increased expression of IL-6 and TNF, which may affect the metabolic function and insulin sensitivity of neighboring muscle tissue,^9–11^ but the impact of IMAT on CVD events is not well understood.

Coronary microvascular dysfunction (CMD), quantified noninvasively using positron emission tomography (PET) as an impaired global coronary flow reserve (CFR <2) with normal myocardial perfusion imaging, is independently associated with elevated BMI^12^ and future risk of heart failure, myocardial infarction and death.^13–18^ It is also associated with residual inflammation and myocardial stiffness independently of conventional CVD risk factors in patients with cardiometabolic disease.^19, 20^ We previously demonstrated an independent inverted J-shaped relationship between BMI and CFR such that in obese patients, CFR decreased linearly with increasing BMI (adjusted p<0.0001).^12^ We found that CMD was prevalent in obese patients, worsened with increasing BMI and was a better discriminator of CVD risk as compared to BMI. Given the limitations of BMI, we sought to investigate the relationship between IMAT, CMD and cardiovascular outcomes. We hypothesized that measures of skeletal muscle quantity and quality are associated with coronary microvascular dysfunction and modify its effect on CVD events independently of obesity.

## Methods

### Study population

The study population (**Supplemental Figure S1**) included consecutive patients undergoing cardiac stress testing with PET/CT at Brigham and Women’s Hospital (Boston, Massachusetts) from 2007 to 2014. The most common indication for PET was the evaluation of chest pain, dyspnea or their combination. Patients with known coronary artery disease (CAD), including a history of previous myocardial infarction or revascularization, heart failure, severe valvular disease, PET evidence of flow-limiting CAD (defined as a summed stress score >2) or abnormal left ventricular ejection fraction (LVEF, <40%) were excluded, as were patients with end-stage disease, including liver, lung or kidney disease, active malignancy or planned bariatric surgery at the time of imaging. Medical history, medication use, laboratory findings, height and weight were ascertained at the time of PET/CT imaging. BMI was calculated as the ratio between weight in kilograms and the square of height in meters. The Chronic Kidney Disease Epidemiology Collaboration equation was used to determine eGFR. The study was approved by the Mass General Brigham Healthcare Institutional Review Board and performed in accordance with institutional guidelines.

### Positron emission tomography imaging

Patients were imaged with a whole-body PET/CT scanner (Discovery RX or STE LightSpeed 64, GE Healthcare, Milwaukee, Wisconsin) using ^82^Rubidium and ^13^N-ammonia as flow radiotracers at rest and following pharmacological stress. Patients were instructed to avoid caffeine or methylxanthine-containing substances 24 hours before the scan. Rest LVEF was obtained from gated myocardial perfusion images processed with commercially available software (Corridor4DM, INVIA Medical Imaging Solutions; Ann Arbor, Michigan). Semiquantitative visual interpretation was performed using a standard 17-segment, 5-point scoring system to determine summed rest, stress and difference scores, reflecting scar, ischemia plus scar, or ischemia, respectively. Absolute global myocardial blood flow (MBF, ml/min/g) was measured at rest and peak vasodilator-induced hyperemia and dynamic images were fitted into a validated two-compartment kinetic model as previously described.^22^ CFR was obtained as the ratio of global stress to rest MBF.

### CT-derived body composition analysis

Thoracic body composition metrics, including cross-sectional areas (in cm^2^) of subcutaneous adipose tissue (SAT), skeletal muscle (SM), and intermuscular adipose tissue (IMAT), were measured at the level of the twelfth vertebra (T12) on non-contrast axial CT images obtained for attenuation correction during standard cardiac PET/CT imaging. Studies with artifacts related to patient positioning or image acquisition precluding soft tissue assessment at this level were excluded (**Supplemental Figure S1**). SAT, SM and IMAT were quantified using semi-automated threshold-based segmentation in 3DSlicer (version 4.10.1, https://www.slicer.org/) by a trained reader (AST) blinded to study details. Tissue-specific Hounsfield units (HU) were used for skeletal muscle (-29 to +150 HU) and adipose tissue (-190 to -30 HU) as previously described.^23, 24^ Boundaries of each compartment were verified with excellent intra-reader class correlations (≥0.997). Randomly selected samples were re-segmented by independent readers (JPM, FJF) blinded to patient details with excellent inter-reader class correlations (≥0.972). All body composition analyses were blinded to clinical, PET and outcomes data.

### Outcomes

Patients were followed over a median 5.8 (Q1-Q3 3.2–7.1) years for the occurrence of a primary endpoint composite of death, hospitalization for nonfatal myocardial infarction (MI) or heart failure (HF). Time to first event was analyzed. Ascertainment of clinical endpoints was determined by blinded committee adjudication of the longitudinal medical record, the Mass General Brigham Healthcare Research Patient Data Registry, the National Death Index, mail surveys, and telephone calls. To be classified as hospitalization for nonfatal MI or HF, discharge with the corresponding primary diagnosis was required, and only events meeting the universal definition of MI^25^ or prespecified criteria for clinical signs, symptoms and escalation of therapy for HF, respectively, were adjudicated as such. All hospitalization events occurred >30 days following PET imaging.

### Statistical analyses

Baseline characteristics are presented as rates with percentages (%) for categorical variables and medians with interquartile ranges (Q1-Q3) for continuous variables. We used the Fisher’s exact test and the Wilcoxon rank-sum test to assess for differences in categorical and continuous baseline characteristics, respectively. CMD was defined as CFR <2, a prognostically important cutpoint associated with worse cardiovascular outcomes in patients undergoing evaluation for suspected CAD,^27, 28^ and approximated the median CFR of the clinical cohort. Spearman’s correlation was used to describe the association between the continuous variables of BMI and thoracic body composition metrics SAT, IMAT and SM. Fatty muscle fraction (FMF, %) was defined as IMAT/(SM+IMAT)*100.

Linear regression was used to assess for independent relationships between CFR and body composition metrics. Candidate variables tested included demographic characteristics, medical history and medication use, laboratory findings and noninvasive imaging parameters, with the most clinically important covariates or significant univariable associations included in the multivariable model. To avoid overfitting, demographic and medical history variables (age, sex, anginal symptoms, hypertension, dyslipidemia, diabetes, tobacco use, family history of premature CAD, BMI >27 kg/m^2^, and estrogen status) were incorporated into multivariable modeling using a validated pretest clinical risk score for diagnosing CAD (with values 0-8, 9-15, and 16-24 indicating low, intermediate, and high pretest risk, respectively), as previously described.^29^ The final multivariable model included the pretest clinical score, nonwhite race, BMI, eGFR<60 ml/min/1.73 m^2^, LVEF, SAT, SM and IMAT or FMF. These variables were also incorporated into a logistic regression model to quantify the association between FMF and CMD.

Cumulative event-free survival curves for the composite endpoint of death or hospitalization for nonfatal MI or HF, were compared across categories of CMD and obesity or IMAT median using the log-rank test. Cox proportional hazards models were used to examine for multivariable-adjusted associations of thoracic body composition, CFR and events. Univariate associations were tested and sequential Cox models controlled for effects of clinically important covariates: Model 1 was adjusted for traditional risk factors, including pretest clinical score, race, eGFR, LVEF and BMI, with sequential addition of body composition metrics and CFR to Models 2 and 3, respectively. Nested models were compared with the likelihood ratio test, and the Akaike information criterion was assessed to avoid overfitting. A linear interaction term for CFR and IMAT or FMF was tested for significance in the final adjusted model. The proportional hazards assumption was confirmed for each model using cumulative martingale residuals.

To further investigate the presence of effect modification between CMD and IMAT, Poisson regression was performed to compute adjusted annualized rates of events across categories of CMD and IMAT median after adjustment for pretest clinical score, SM and SAT areas. Model fit was assessed with the goodness-of-fit χ^2^ test, with a nonsignificant result indicating adequate fit. A p-value of <0.05 was considered to indicate statistical significance and all tests were two-sided. Statistical analyses were performed using SAS (version 9.4, SAS Institute Inc., Cary, NC) and SPSS (version 28.0, IBM Corp, Armonk, NY).

## Results

### Baseline Characteristics

The distribution of baseline characteristics is shown in **Table 1**. The median age of patients in the overall cohort was 62.6 (53.7-71.6) years, 69.8% were women and 46.2% were nonwhite. Approximately 45.9% of patients were obese (BMI 30-61 kg/m^2^), with 21.7% of the cohort classified as having class I obesity (BMI 30-34.9 kg/m^2^), 13.1% as class II obesity (BMI 35-39.9 kg/m^2^) and 11.1% as class III obesity (BMI ≥40 kg/m^2^). Hypertension and dyslipidemia were prevalent comorbidities present in 72.6% and 58.4% of patients, respectively, and 25.6% of patients had diabetes. Median LVEF was 63% (57-70) and 43.0% of patients had CMD, with median coronary flow reserve of 2.1 (1.7-2.6) reflecting median peak stress and rest myocardial blood flow values of 2.3 (1.8-2.9) and 1.1 (0.8-1.4) ml/min/g, respectively. Compared with the nonobese, obese patients had higher median indices of SAT (248.7 vs. 101.5 cm^2^), SM (95.6 vs. 78.9 cm^2^) and IMAT (20.4 vs. 6.5 cm^2^, p<0.001 for all), and similar distributions of CFR. BMI correlated highly with SAT and IMAT (r=0.84 and 0.71, respectively, p<0.001) and moderately with SM (r=0.52, p<0.001) (**Figure 1, Supplemental Figure S2**). Representative patients of similar demographics and BMI with variable thoracic body composition metrics and CFR values are shown in **Figure 2**.

**Figure 1.**
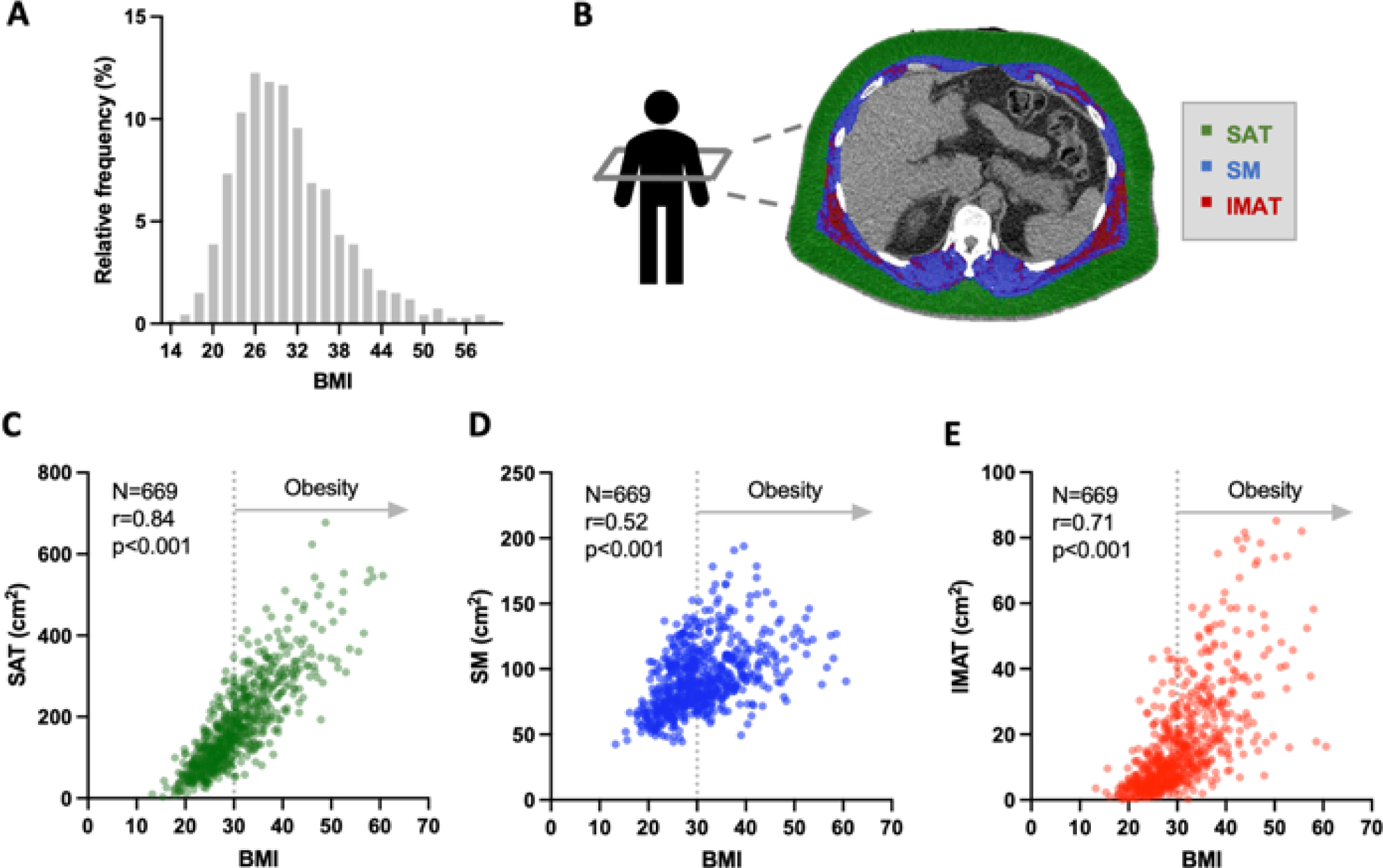
Thoracic body composition compartments and relationship to body mass index, BMI. (A) Distribution of BMI in the study population. (B) Example of CT slice selection at the twelfth thoracic vertebra (T12) level and segmentation of subcutaneous adipose tissue (SAT, green), skeletal muscle (SM, blue) and intermuscular adipose tissue (IMAT, red) areas. Relationship between BMI and SAT (C), SM (D), and IMAT (E).

**Figure 2.**
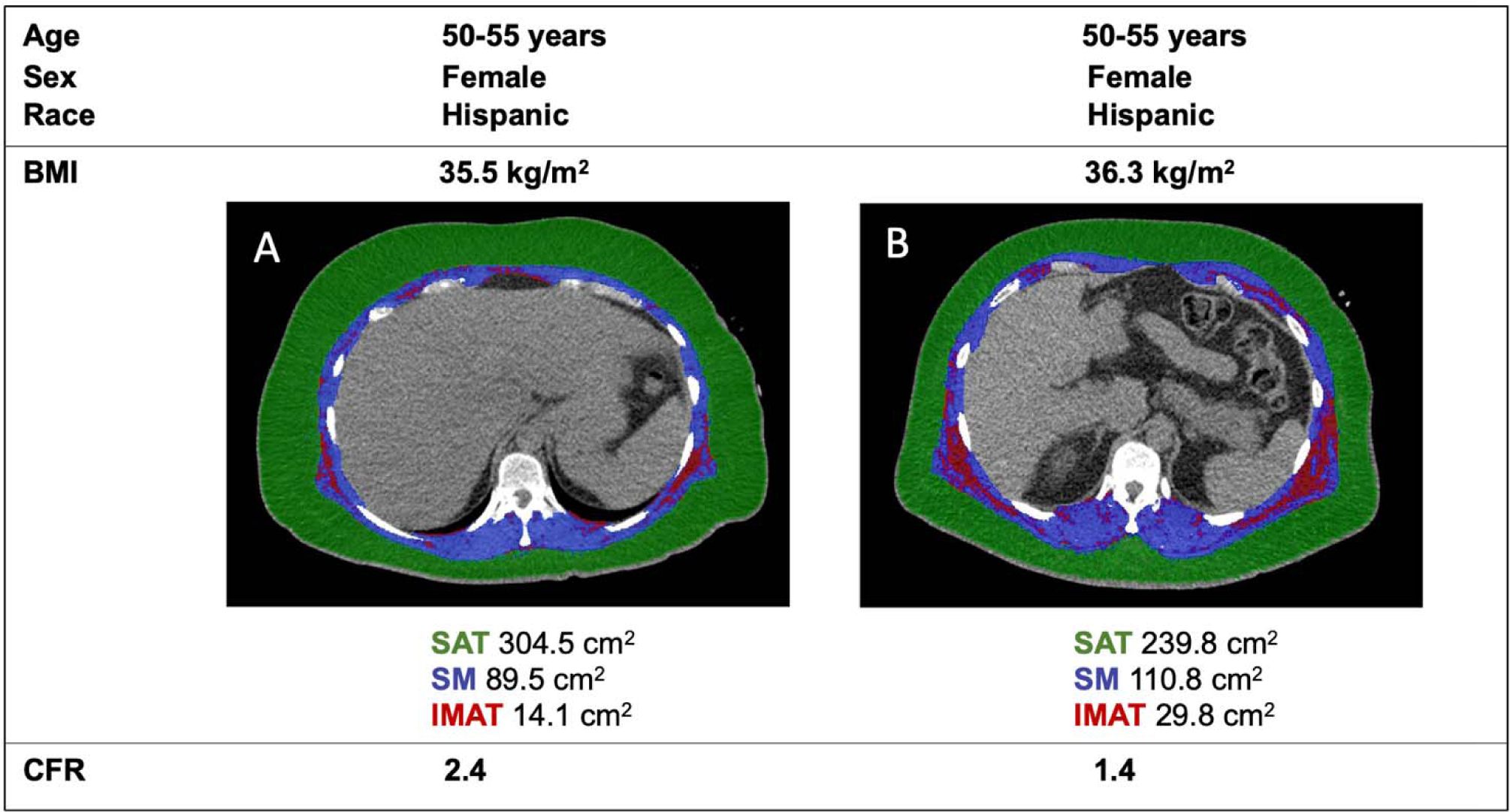
Characterization of thoracic body composition at the twelfth thoracic vertebra (T12) level in representative patients (A, B) of similar age, sex, race and BMI with normal renal function, LVEF and myocardial perfusion. Relative to patient (A), patient (B) demonstrated decreased SAT and increased SM and IMAT areas, also with CFR <2, consistent with CMD. BMI: body mass index; LVEF: left ventricular ejection fraction; SAT: subcutaneous adipose tissue; SM: skeletal muscle; IMAT: intermuscular adipose tissue; CFR: coronary flow reserve; CMD: coronary microvascular dysfunction

**Table 1.** Baseline Characteristics of the Study Population.

| Characteristic | Overall<br>(N=669) | Obese* |  | P-value† |
| --- | --- | --- | --- | --- |
|  |  | No (n=362) | Yes (n=307) |  |
| Demographics |  |  |  |  |
| Age (years) | 62.6 (53.7-71.6) | 64.9 (55.1-74.2) | 60.1 (52.1-67.1) | <0.001 |
| Female (%) | 467 (69.8%) | 236 (65.2%) | 231 (75.2%) | 0.005 |
| Nonwhite race (%) | 309 (46.2%) | 146 (40.3%) | 163 (53.1%) | 0.001 |
| Body mass index (kg/m<sup>2</sup>) | 29.2 (25.1-34.7) | 25.6 (23.1-27.8) | 35.4 (32.3-39.8) | <0.001 |
| Pretest clinical score‡ | 11 (8-13) | 11.0 (8.0-13.8) | 10.0 (8.0-14.0) | 0.56 |
| Medical history |  |  |  |  |
| Hypertension (%) | 486 (72.6%) | 231 (63.8%) | 255 (83.1%) | <0.001 |
| Dyslipidemia (%) | 391 (58.4%) | 202 (55.8%) | 189 (61.6%) | 0.14 |
| Diabetes (%) | 171 (25.6%) | 57 (15.7%) | 114 (37.1%) | <0.001 |
| Family history of CAD (%) | 195 (29.1%) | 89 (24.6%) | 106 (34.5%) | 0.006 |
| Current tobacco use (%) | 56 (8.4%) | 31 (8.56%) | 25 (8.14%) | 0.89 |
| Medication use |  |  |  |  |
| Aspirin (%) | 353 (52.8%) | 199 (55.0%) | 154 (50.2%) | 0.24 |
| Beta-blocker (%) | 289 (43.2%) | 145 (40.1%) | 144 (46.9%) | 0.09 |
| Angiotensin inhibitor (%) | 197 (29.4%) | 88 (24.3%) | 109 (35.5%) | 0.002 |
| Statin (%) | 343 (51.3%) | 177 (48.9%) | 166 (54.1%) | 0.19 |
| Oral hypoglycemic (%) | 80 (12%) | 24 (6.63%) | 56 (18.2%) | <0.001 |
| Insulin (%) | 71 (10.6%) | 18 (4.97%) | 53 (17.3%) | <0.001 |
| Laboratory values |  |  |  |  |
| Creatinine (mg/dL) | 0.83 (0.72-0.96) | 0.8 (0.7-1.0) | 0.8 (0.7-0.9) | 0.24 |
| eGFR (ml/min/1.73 m<sup>2</sup>) | 84.3 (70.1-96.7) | 82.1 (70.4-93.8) | 87.1 (70.2-98.6) | 0.02 |
| CT body composition measures |  |  |  |  |
| Subcutaneous adipose tissue (SAT, cm<sup>2</sup>) | 155.4 (92.7-246.8) | 101.5 (69.3-139.9) | 248.7 (187.9-317.8) | <0.001 |
| Skeletal muscle (SM, cm<sup>2</sup>) | 87.7 (73.5-106.2) | 78.9 (67.8-98.7) | 95.6 (82.4-114.5) | <0.001 |
| Intermuscular adipose tissue (IMAT, cm<sup>2</sup>) | 11.5 (5.6-22.7) | 6.5 (4.0-11.3) | 20.4 (13.2-34.2) | <0.001 |
| Fatty muscle fraction (FMF, %) | 11.6 (6.3-19.1) | 7.5 (4.6-12.7) | 18.0 (11.7-25.2) | <0.001 |
| PET imaging parameters |  |  |  |  |
| Left ventricular ejection fraction (%) | 63 (57-69.5) | 63.0 (57.2-70.0) | 63.0 (56.0-68.0) | 0.46 |
| Rest myocardial blood flow (mL/min/g) | 1.1 (0.8-1.4) | 1.1 (0.9-1.5) | 1.0 (0.8-1.3) | <0.001 |
| Stress myocardial blood flow (mL/min/g) | 2.3 (1.8-2.9) | 2.5 (2.0-3.1) | 2.1 (1.6-2.6) | <0.001 |
| Coronary flow reserve | 2.1 (1.7-2.6) | 2.1 (1.7-2.6) | 2.1 (1.6-2.5) | 0.07 |

Continuous variables are presented as medians (Q1-Q3) and categorical variables as (%).
\*Obese is defined as body mass index $\geq 30$ .
†The P-value is for the comparison between obese groups and is based on the Fisher exact test for categorical variables and the Wilcoxon rank sum test for continuous variables.

### Association Between Thoracic Body Composition and Coronary Flow Reserve

In univariable linear regression analysis, increasing BMI was inversely associated with CFR [β(se)=-0.07(0.03), p=0.04 for +10 kg/m^2^ BMI], as was increasing SAT and IMAT [β(se)=-0.007(0.002) and -0.053(0.016), p=0.003 and 0.001 for +10 cm^2^ SAT and IMAT areas, respectively] (**Table 2**). After adjustment for clinical covariates and body composition metrics, decreasing SM and increasing IMAT, but not BMI or SAT, remained independently associated with worse CFR [β(se)=0.027(0.013) and -0.047(0.023), adjusted p=0.03 and 0.04 for +10 cm^2^ SM and IMAT areas, respectively] (**Table 2**). Findings were confirmed using FMF [β(se)=-0.008(0.004), adjusted p=0.02 for +1% FMF]. In univariable and multivariable-adjusted logistic regression analyses, every 1% increase in FMF was associated with a 2% increased odds of CMD [OR 1.02 (1.01-1.04), unadjusted p=0.008 and adjusted p=0.04] defined as CFR <2.

**Table 2.** Univariable and Multivariable-adjusted Associations of Thoracic Body Composition and CFR.

|  | Univariable |  | Multivariable* |  |
| --- | --- | --- | --- | --- |
| | $\beta$ (se) | P-value | $\beta$ (se) | P-value |
| Pretest clinical score | -0.016 (0.006) | 0.01 | -0.017 (0.006) | 0.008 |
| Nonwhite race | 0.041 (0.051) | 0.42 | 0.039 (0.052) | 0.46 |
| eGFR<60 ml/min/1.73 m<sup>2</sup> | -0.020 (0.085) | 0.81 | 0.063 (0.086) | 0.47 |
| LVEF (+10%) | -0.058 (0.028) | 0.04 | -0.040 (0.029) | 0.17 |
| BMI (+10 kg/m<sup>2</sup>) | -0.069 (0.033) | 0.04 | 0.005 (0.072) | 0.94 |
| SAT (+10 cm<sup>2</sup>) | -0.007 (0.002) | 0.003 | -0.005 (0.005) | 0.32 |
| SM (+10 cm<sup>2</sup>) | 0.014 (0.010) | 0.18 | 0.027 (0.013) | 0.03 |
| IMAT (+10 cm<sup>2</sup>) | -0.053 (0.016) | 0.001 | -0.047 (0.023) | 0.04 |
| FMF<sup>^</sup> (+1%) | -0.011 (0.003) | <0.0001 | -0.008 (0.004) | 0.02 |

$\beta$ estimates with standard error (se) are listed.
\*Adjusted for pretest clinical score, nonwhite race, eGFR <60 ml/min/1.73 m<sup>2</sup>, LVEF, BMI, SAT, SM and IMAT or FMF.
BMI: body mass index; SAT: subcutaneous adipose tissue; SM: skeletal muscle; IMAT: intermuscular adipose tissue; FMF: fatty muscle fraction, IMAT/(SM+IMAT)\*100
<sup>^</sup>Every 1% increase in FMF was associated with a 2% increased odds of CMD, defined as CFR <2 [OR 1.02 (1.01-1.04), unadjusted p=0.008 and adjusted p=0.04]

### Thoracic Body Composition, Coronary Flow Reserve and Adverse Events

Over a median follow-up of 5.8 (3.2-7.1) years, 98 patients met the composite endpoint of death and hospitalization for MI or HF, including 54 deaths (**Supplemental Table S1**). In univariable analysis, CFR and IMAT were significantly associated with major adverse events (HR 1.81, 95%CI 1.28-2.56 per -1U CFR, p=0.001 and HR 1.21, 95%CI 1.09-1.35 per +10 cm^2^ IMAT, p<0.001, respectively) (**Table 3**). The addition of body composition metrics SAT, SM and IMAT into a multivariable model of clinically important covariates, including pretest clinical score, race, BMI, eGFR and LVEF led to improvement in model statistics (Global Chi-square 65.4 vs 26.8 for Model 2 vs 1, respectively, p<0.001), as did the sequential addition of CFR (Global Chi-square 74.5 vs 65.4 for Model 3 vs 2, respectively, p<0.001p<0.001 for Model 3 vs 2) (**Table 3**). In the final adjusted model, both lower CFR and higher IMAT were associated with increased events [HR 1.78 (1.23-2.58) per -1U CFR and 1.53 (1.30-1.80) per +10 cm^2^ IMAT, adjusted p=0.002 and p≤0.0001, respectively], while higher SM and SAT were protective [HR 0.89 (0.81-0.97) per +10 cm^2^ SM and 0.94 (0.91-0.98) per +10 cm^2^ SAT, adjusted p=0.01 and 0.003, respectively]. There was a significant interaction between CFR and IMAT such that patients with both CMD and fatty muscle demonstrated the highest risk of events (adjusted p for interaction=0.02). Every 1% increase in fatty muscle fraction conferred an independent 7% increased risk of major adverse events [HR 1.07 (1.04-1.09), adjusted p<0.001], and modified the effect of CMD on outcomes (adjusted p for interaction=0.04).

**Table 3.**
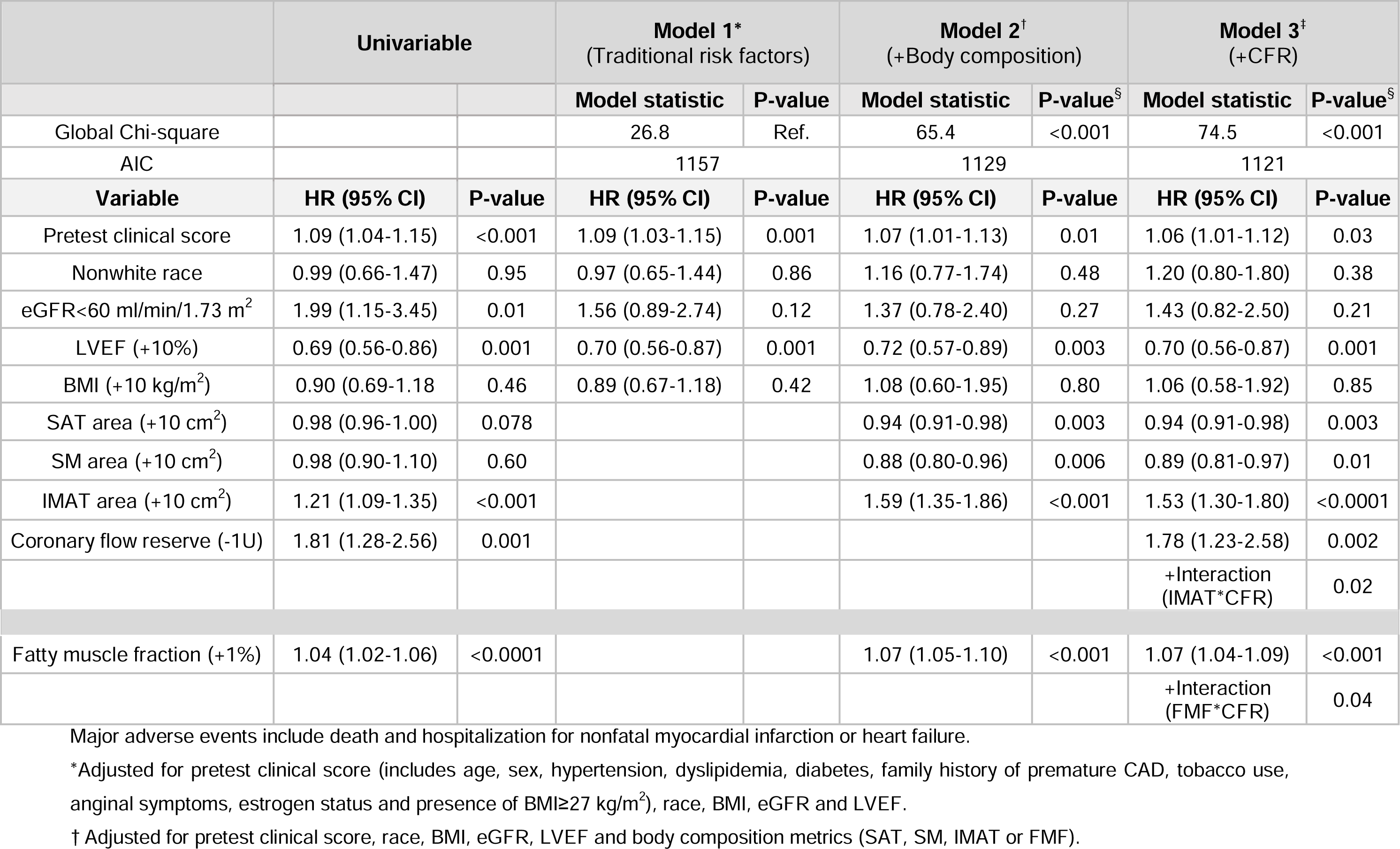
Multivariable-adjusted Associations of Thoracic Body Composition and CFR with Major Adverse Events.

|  | Univariable |  | Model 1*<br>(Traditional risk factors) |  | Model 2†<br>(+Body composition) |  | Model 3‡<br>(+CFR) |  |
| --- | --- | --- | --- | --- | --- | --- | --- | --- |
|  |  |  | Model statistic | P-value | Model statistic | P-value<sup>§</sup> | Model statistic | P-value<sup>§</sup> |
| Global Chi-square |  |  | 26.8 | Ref. | 65.4 | <0.001 | 74.5 | <0.001 |
| AIC |  |  | 1157 |  | 1129 |  | 1121 |  |
| Variable | HR (95% CI) | P-value | HR (95% CI) | P-value | HR (95% CI) | P-value | HR (95% CI) | P-value |
| Pretest clinical score | 1.09 (1.04-1.15) | <0.001 | 1.09 (1.03-1.15) | 0.001 | 1.07 (1.01-1.13) | 0.01 | 1.06 (1.01-1.12) | 0.03 |
| Nonwhite race | 0.99 (0.66-1.47) | 0.95 | 0.97 (0.65-1.44) | 0.86 | 1.16 (0.77-1.74) | 0.48 | 1.20 (0.80-1.80) | 0.38 |
| eGFR<60 ml/min/1.73 m<sup>2</sup> | 1.99 (1.15-3.45) | 0.01 | 1.56 (0.89-2.74) | 0.12 | 1.37 (0.78-2.40) | 0.27 | 1.43 (0.82-2.50) | 0.21 |
| LVEF (+10%) | 0.69 (0.56-0.86) | 0.001 | 0.70 (0.56-0.87) | 0.001 | 0.72 (0.57-0.89) | 0.003 | 0.70 (0.56-0.87) | 0.001 |
| BMI (+10 kg/m<sup>2</sup>) | 0.90 (0.69-1.18) | 0.46 | 0.89 (0.67-1.18) | 0.42 | 1.08 (0.60-1.95) | 0.80 | 1.06 (0.58-1.92) | 0.85 |
| SAT area (+10 cm<sup>2</sup>) | 0.98 (0.96-1.00) | 0.078 |  |  | 0.94 (0.91-0.98) | 0.003 | 0.94 (0.91-0.98) | 0.003 |
| SM area (+10 cm<sup>2</sup>) | 0.98 (0.90-1.10) | 0.60 |  |  | 0.88 (0.80-0.96) | 0.006 | 0.89 (0.81-0.97) | 0.01 |
| IMAT area (+10 cm<sup>2</sup>) | 1.21 (1.09-1.35) | <0.001 |  |  | 1.59 (1.35-1.86) | <0.001 | 1.53 (1.30-1.80) | <0.0001 |
| Coronary flow reserve (-1U) | 1.81 (1.28-2.56) | 0.001 |  |  |  |  | 1.78 (1.23-2.58) | 0.002 |
|  |  |  |  |  |  |  | +Interaction<br>(IMAT*CFR) | 0.02 |
| Fatty muscle fraction (+1%) | 1.04 (1.02-1.06) | <0.0001 |  |  | 1.07 (1.05-1.10) | <0.001 | 1.07 (1.04-1.09) | <0.001 |
|  |  |  |  |  |  |  | +Interaction<br>(FMF*CFR) | 0.04 |

\*Adjusted for pretest clinical score (includes age, sex, hypertension, dyslipidemia, diabetes, family history of premature CAD, tobacco use, anginal symptoms, estrogen status and presence of BMI ≥ 27 kg/m<sup>2</sup>), race, BMI, eGFR and LVEF.
† Adjusted for pretest clinical score, race, BMI, eGFR, LVEF and body composition metrics (SAT, SM, IMAT or FMF).

### Obesity, Intermuscular Adiposity, Coronary Microvascular Dysfunction and Outcomes

To visualize the impact of effect modification of intermuscular adipose tissue on coronary microvascular dysfunction and cardiovascular events, we stratified results by CMD and obesity (**Figures 3A, 3B**) or IMAT median (**Figures 3C, 3D**). In adjusted analysis, patients with CMD with or without obesity experienced the highest cumulative rate of events (adjusted p<0.001, **Figure 3B**). Elevated BMI did not further stratify risk among patients with CMD (**Figures 3B, 4A**). In contrast, only those patients with both CMD and high IMAT experienced the highest rate of adverse events (adjusted p<0.001, **Figures 3D, 4B**), with an adjusted annualized rate of events of 5.1% (adjusted p=0.02).

**Figure 3.**
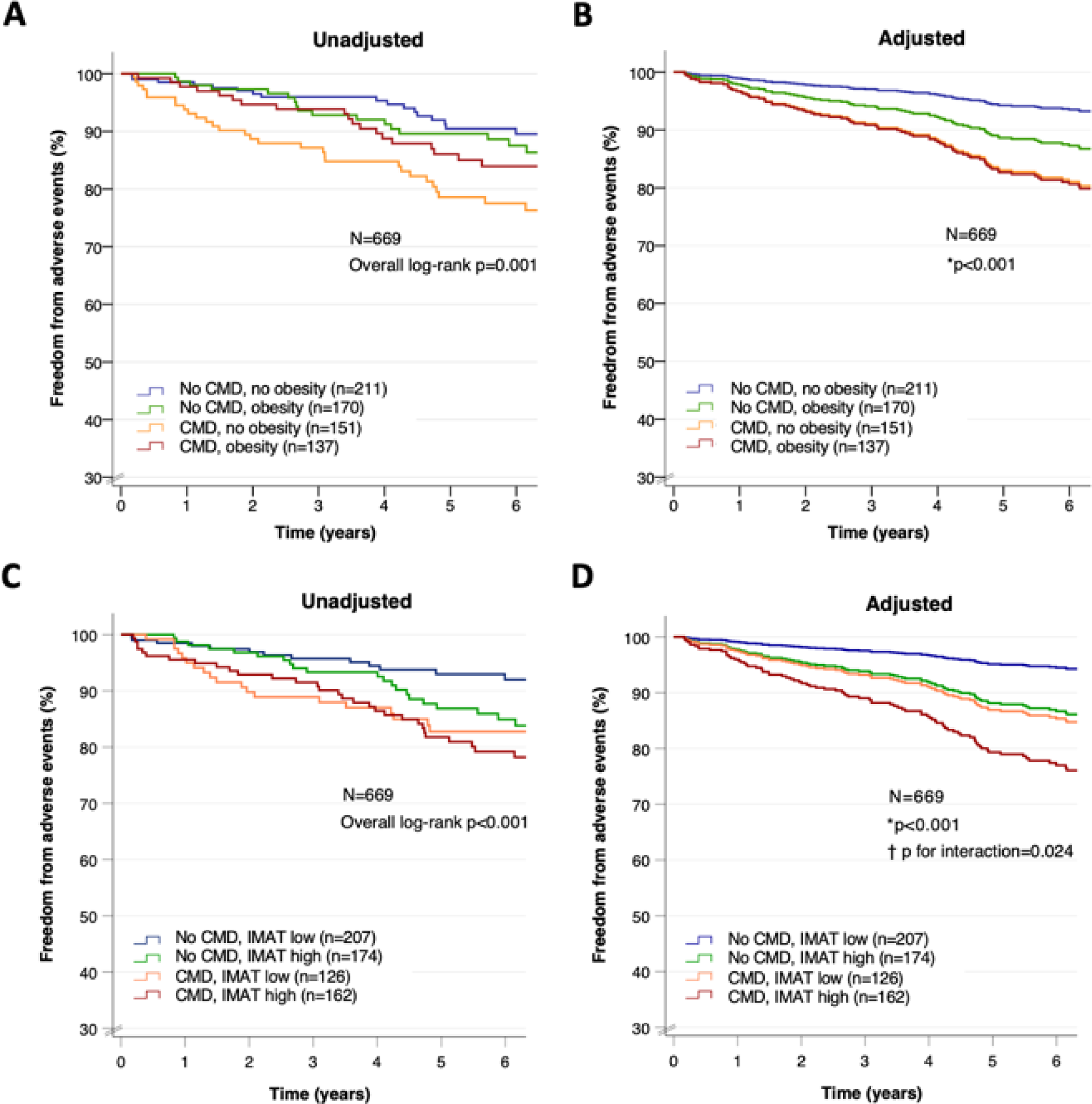
Unadjusted (A, C) and adjusted (B, D) freedom from major adverse events by CMD and obesity (A, B) or IMAT median (C, D). Major adverse events include death and hospitalization for nonfatal myocardial infarction or heart failure. CMD: coronary microvascular dysfunction, coronary flow reserve <2; obesity, body mass index ≥30; IMAT: intermuscular adipose tissue, high ≥11.5 cm^2^ *Adjusted for pretest clinical score, race, estimated glomerular filtration rate <60, left ventricular ejection fraction, skeletal muscle and subcutaneous adipose tissue areas ^†^ p-value refers to interaction between coronary flow reserve and IMAT continuous variables

**Figure 4.**
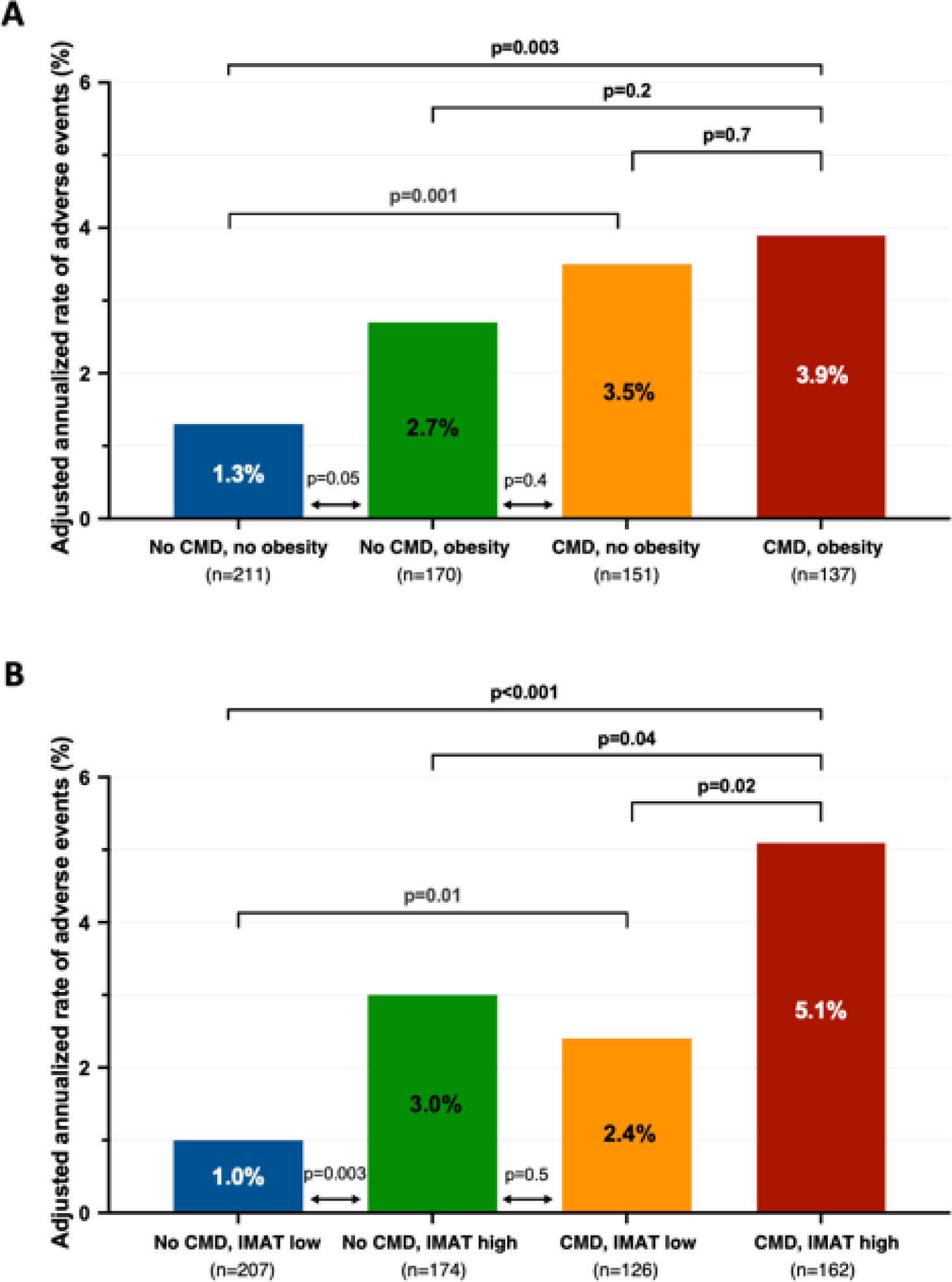
Adjusted annualized rate of major adverse events by CMD and obesity (A) or IMAT median (B). Major adverse events include death and hospitalization for nonfatal myocardial infarction or heart failure. Poisson regression was adjusted for pretest clinical score, skeletal muscle and subcutaneous adipose tissue areas. CMD: coronary microvascular dysfunction, coronary flow reserve <2; obesity, body mass index ≥30; IMAT: intermuscular adipose tissue, high ≥11.5 cm^2^

## Discussion

In this work, we leveraged advances in noninvasive imaging to demonstrate a novel relationship between IMAT, CMD and cardiovascular outcomes. First, we found that lower skeletal muscle quantity and quality was associated with coronary microvascular dysfunction, independently of conventional risk factors. Every 1% increase in fatty muscle fraction conferred an independent 2% increased odds of CMD. Second, in adjusted analyses for outcomes, we showed that both higher IMAT and lower CFR were associated with increased risk of CVD events, while higher SM and SAT were protective. Every 1% increase in fatty muscle fraction conferred a 7% increased risk of CVD events. Third, the presence of elevated IMAT, not BMI, modified the effect of CMD on outcomes such that patients with both CMD and fatty muscle, not obesity, demonstrated the highest risk of CVD events. Our data support that thoracic skeletal muscle quantity *and* quality are tied to coronary microvascular function and may help to identify at-risk cardiometabolic phenotypes beyond traditional measures of adiposity.

These findings may hold special relevance for the common clinical demographic represented here, namely women and patients with ischemia and no obstructive CAD (INOCA). Obesity-related CVD risk is not well-captured by BMI, especially in women.^30^ While increased thoracic IMAT was independently associated with worse CVD events, higher thoracic SM and, to a lesser extent, SAT demonstrated a protective effect. This underscores that in INOCA patients, assessments of BMI and traditional adiposity – which largely reflect lower-risk SAT – are likely to be *insufficient* to appropriately stratify CVD risk. Our findings are clinically relevant and may help to better discern CVD risk among otherwise similar patients, especially younger, female individuals with fewer traditional risk factors and high prevalence of overweight/obesity. **Figures 2A and 2B** demonstrate thoracic body composition results in 2 middle-aged Hispanic women with class II obesity. Despite similar age and BMI, normal renal function, LVEF and myocardial perfusion, patient (B) has increased IMAT and decreased CFR as compared to patient (A), and thus may be at a significantly increased risk of CVD events.

Described not long ago, IMAT reflects an ectopic adipose tissue depot that is interspersed among skeletal muscle fibers residing within the boundary of the muscle fascia.^8^ Considered a marker of overall muscle quality, it has been associated with systemic inflammation, insulin resistance, the metabolic syndrome, and coronary artery calcification in a manner comparable to visceral adipose tissue.^11, 31, 32^ We recently reported on the independent associations between residual inflammation, abnormal coronary flow reserve and impaired myocardial strain in a cohort of clinical trial patients with cardiometabolic disease and otherwise well-controlled CVD risk factors and preserved cardiac function.^19^ We found that individuals with impaired CFR demonstrated the strongest association between markers of heart failure and inflammation, building on prior work identifying abnormal global CFR and CMD as robust markers of future risk of heart failure hospitalization in patients with preserved LVEF.^14–16^ HFpEF is prevalent in obese patients and often preceded by exercise intolerance, which may reflect limitations in skeletal muscle related to functional and structural impairments also common to the myocardium, including reduced oxidative capacity, microvascular dysfunction, capillary rarefaction and fibrosis.^33–35^

The expansion of IMAT is thought to promote an adverse muscle phenotype associated with metabolic derangements, including impaired bioenergetics, mitochondrial dysfunction, and increased catabolism with loss of lean muscle mass.^8^ Emerging data suggest that the human IMAT secretome is highly immunogenic and inflammatory relative to that of SAT or even VAT depots in individuals with obesity, and may alter immune cell trafficking to neighboring muscle in a manner that may be critical to regional glucose homeostasis.^11^ Taken together, these findings support the hypothesis that common systemic factors involving inflammation and altered glucose metabolism may promote parallel insults to myocardial and skeletal muscle beds, which may manifest as microvascular dysfunction and intramuscular fat infiltration, respectively, and ultimately contribute to adverse CVD events, including HFpEF outcomes.

Our study has important limitations including its observational, single-center design. Despite the use of multivariable-adjusted models and confirmatory testing showing robust data results, residual confounding may persist. Although we leveraged low-dose CTs, which were routinely obtained with clinical cardiac PET studies for the purpose of attenuation correction, we were able to demonstrate excellent segmentation quality assurance, and the ability to obtain concurrent imaging in these patients represents an important strength of this study. Because these were thoracic studies, we focused our cross-sectional assessments at the level of T12 for thoracic body composition analysis. As such, results may differ when compared to abdominal body composition results obtained from the lumbar spine level,^36^ and visceral adipose tissue could not be accurately quantified (although we would expect it to have a less important role in this female-predominant cohort since women have significantly lower visceral and higher subcutaneous adiposity as compared with men^30^). Since cardiac PET is performed using vasodilator stress, no exercise testing results were available. As only patients with normal visual myocardial perfusion and no evidence of flow-limiting CAD were included, the patient cohort was predominantly female, although findings are generalizable to an INOCA population. Relative to less accurate or more cumbersome methods for body composition analysis,^37^ opportunistic CT used in this manner may provide personalized prognostic information without added cost or radiation exposure, and is poised to grow with rapid developments in artificial intelligence and machine learning. Future studies assessing the impact of therapeutic strategies, such as supervised exercise and nutritional, medical and/or surgical weight loss interventions on skeletal muscle quality and CMD outcomes are warranted.

## Conclusion

In patients without flow-limiting CAD, increased intermuscular fat is associated with CMD and adverse cardiovascular outcomes independently of BMI and conventional risk factors. The presence of CMD and skeletal muscle fat infiltration identified a novel at-risk cardiometabolic phenotype.

## Clinical Perspective

### What is new?

- Intermuscular fat infiltration is associated with coronary microvascular dysfunction (CMD) and adverse cardiovascular outcomes independently of BMI and conventional risk factors.
- Every 1% increase in thoracic fatty muscle fraction conferred an independent 2% increased odds of CMD and a 7% increased risk of adverse cardiovascular events.
- The presence of both coronary microvascular dysfunction and skeletal muscle fat infiltration identified a novel at-risk cardiometabolic phenotype prevalent in patients with ischemia and no obstructive CAD (INOCA).

### What are the clinical implications?

- Obesity-related cardiovascular risk is not well-captured by BMI, especially in those with few traditional risk factors and greater subcutaneous adiposity.
- Skeletal muscle quantity *and* quality are tied to coronary microvascular function and may help to identify cardiometabolic risk beyond traditional measures of adiposity, especially in women.

## Data Availability

All data produced in the present study are under the oversight of Mass General Brigham Healthcare institutional guidelines.

## Supplemental Materials

**Table S1:**
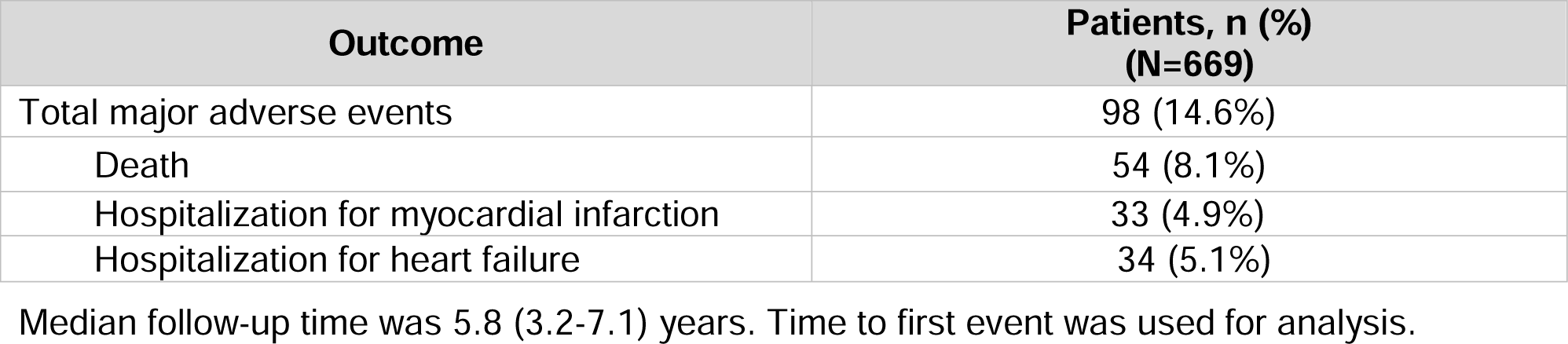
Patients Meeting the Clinical Endpoint.

| **Outcome** | **Patients, n (%)**<br>**(N=669)** |
| --- | --- |
| Total major adverse events | 98 (14.6%) |
| Death | 54 (8.1%) |
| Hospitalization for myocardial infarction | 33 (4.9%) |
| Hospitalization for heart failure | 34 (5.1%) |

Median follow-up time was 5.8 (3.2-7.1) years. Time to first event was used for analysis.

**Figure S1.**
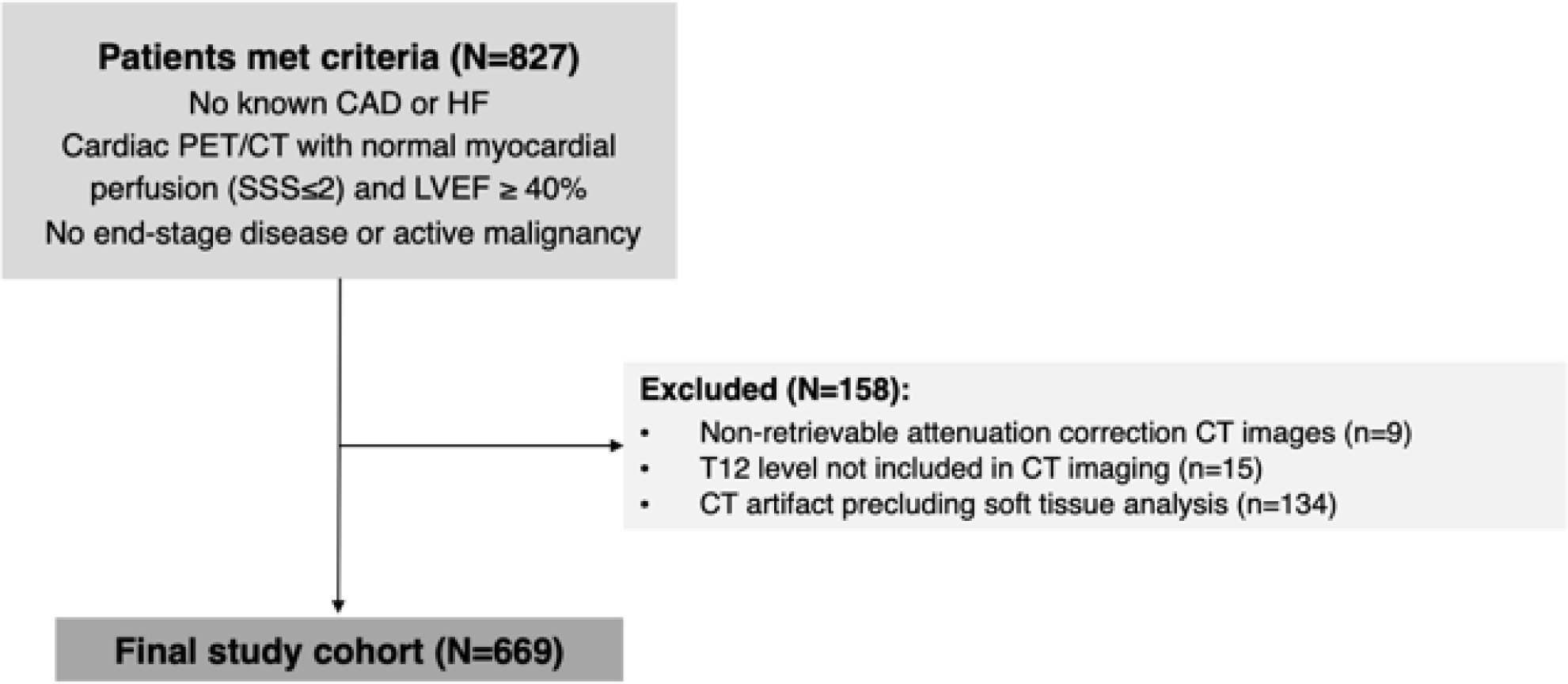
Study flowchart. CAD: coronary artery disease; HF: heart failure; PET: positron emission tomography; CT: computed tomography; SSS: summed stress score; LVEF: left ventricular ejection fraction; T12: twelfth thoracic vertebra

**Figure S2.**
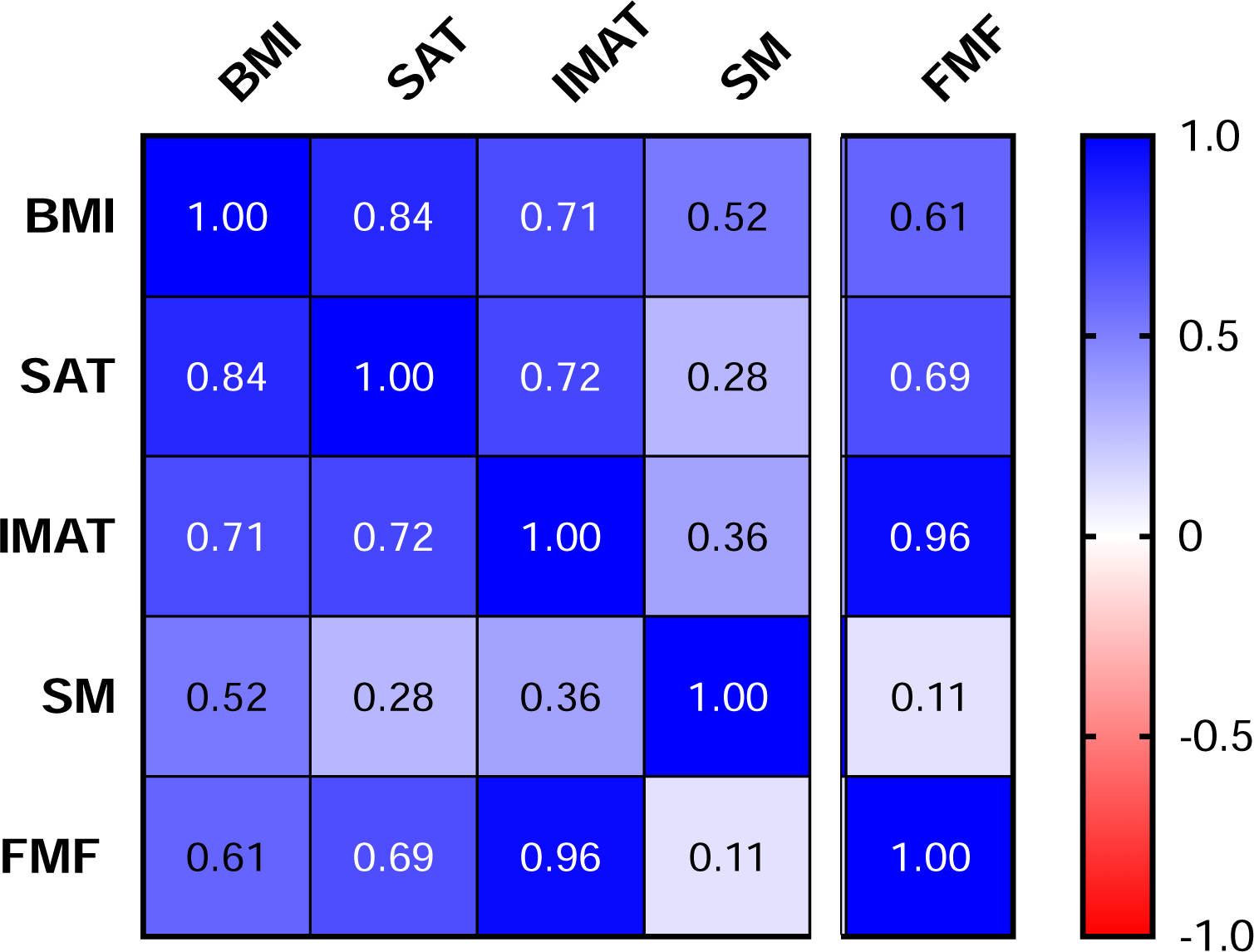
Spearman’s correlations between measures of BMI and thoracic body composition, p<0.005 for all. BMI: body mass index; SAT: subcutaneous adipose tissue; IMAT: intermuscular adipose tissue; SM: skeletal muscle; FMF: fatty muscle fraction, IMAT/(SM+IMAT)*100

